# Emergence of SARS-CoV-2 Omicron Variant JN.1 in Tamil Nadu, India - Clinical Characteristics and Novel Mutations

**DOI:** 10.1101/2024.04.16.24305882

**Authors:** Sivaprakasam T. Selvavinayagam, Sathish Sankar, Yean K. Yong, Amudhan Murugesan, Suvaiyarasan Suvaithenamudhan, Kannan Hemashree, Manivannan Rajeshkumar, Anandhazhvar Kumaresan, Ramendra P. Pandey, Saravanan Shanmugam, Parthiban Arthydevi, Masilamani S. Kumar, Natarajan Gopalan, Meganathan Kannan, Narayanaiah Cheedarla, Hong Y. Tan, Ying Zhang, Marie Larsson, Pachamuthu Balakrishnan, Vijayakumar Velu, Siddappa N. Byrareddy, Esaki M. Shankar, Sivadoss Raju

## Abstract

In December 2023, we observed a notable shift in the COVID-19 landscape, when the JN.1 emerged as a predominant SARS-CoV-2 variant with a 95% incidence. We characterized the clinical profile, and genetic changes in JN.1, an emerging SARS-CoV-2 variant of interest. Whole genome sequencing was performed on SARS-CoV-2 positive samples, followed by sequence analysis. Mutations within the spike protein sequences were analyzed and compared with the previous lineages and sublineages of SARS-CoV-2, to identify the potential impact of these unique mutations on protein structure and possible functionality. Several unique and dynamic mutations were identified herein. Our data provides key insights into the emergence of newer variants of SARS-CoV-2 in our region and highlights the need for robust and sustained genomic surveillance of SARS-CoV-2.

## INTRODUCTION

The emergence of the novel coronavirus, SARS-CoV-2, in December 2019, has prompted an urgent need to investigate the evolutionary dynamics of the virus, globally. Since their initial identification, these viruses have constantly undergone mutations, leading to the emergence of several ‘variants of concern’ due to evolutionary dynamics [1]. Viral mutations inherent to the replication process have led to the development of diverse variants characterized by distinct transmissibility and resistance profiles. Since the SARS-CoV-2 pandemic, the virus emerged with five variants of concern (Alpha, Beta, Gamma, Delta, Omicron). Among these, the Omicron variant (B.1.1.529) exhibited more than 50 characteristic mutations in different motifs of the spike protein [2]. The virus along with its subsequent concerning sublineages and variants, evolved with enhanced transmissibility, infectivity and immune evasion mechanism. Notable subvariants, including BA.2, BA.5, and XBB were of particular interest due to their rapid spread across the globe and evasion from neutralizing and monoclonal antibodies [3, 4]. Specific genetic changes within the virus have conferred selective advantages, augmenting its ability to propagate and evade host immune responses, thereby presenting formidable challenges to containment and treatment strategies [5].

These dynamic and ever-evolving viral mutations underscore the imperative necessity for consistent monitoring and use of adaptive methodologies, such as whole-genome sequencing. The ongoing surveillance of viral genomes facilitates the prompt identification of emerging variants, playing a pivotal role in devising public health measures and targeted interventions. Furthermore, a profound understanding of the genetic alterations steering the viral behaviour becomes instrumental in developing effective treatments and vaccines, ensuring a resilient response to the constantly evolving viral threats [6]. The impact of mutations on the protein function is now studied through *in silico* methods [7, 8]. The difference in protein structures induced by mutations resulting in different lineages during the viral evolutionary process [9]. Here, we surveyed the population for SARS-CoV-2 Omicron subvariant JN.1 between November and December 2023 as a part of the state’s public health genomic surveillance investigation, an ongoing programme of the Directorate of Public Health and Preventive Medicine, Chennai, India since September 2021.

## MATERIALS AND METHODS

### Study design

The study was part of the SARS-CoV-2 genomic surveillance by the State Public Health Laboratory (SPHL), Directorate of Public Health and Preventive Medicine, Chennai, India. COVIDLJ19 diagnosis was based on clinical and laboratory tests using nasopharyngeal and oropharyngeal swabs as per the guidelines of the Centers for Disease Control and Prevention (CDC), Atlanta, USA. Written informed consent was obtained before sampling. Patients’ demographic details and clinical presentations were collected, including underlying comorbidities, vaccination history, disease progression, and outcome. Of the 471 COVID-19- positive samples reported in Tamil Nadu between November and December 2023, 92 samples with a cycle threshold (Ct) value of <25 were chosen for whole genome sequencing. RNA extraction was carried out using MagMAX Viral/Pathogen II Nucleic Acid isolation kit (Thermo Fisher Scientific, USA) and tested for SARS-CoV-2 using TaqPath COVID-19 RT-PCR kit (Thermo Fisher Scientific, USA) according to the manufacturer’s instructions. The RNA elutes were stored at -80°C until further testing.

### Ethics approval

The study was approved (EC No. 03092021) by the Institutional Ethics Committee of the Madras Medical College and Hospital, Chennai. The clinical classification was based on the Clinical Guidance for Management of Adult COVIDLJ19 Patients by the Ministry of Health and Family Welfare, Government of India (January 2022).

### Whole genome sequencing

Complementary DNA (cDNA) was prepared from RNA elutes using the SuperScript VILO cDNA Synthesis kit (Invitrogen, Thermo Fisher Scientific, USA) as per the manufacturer’s instructions. Among 471 SARS-CoV-2 positive samples reported during the study period, whole genome sequencing was carried out on 66 of 92 samples with cycle threshold (Ct) values <25 as per the World Health Organization (WHO) criterion. Ion AmpliSeq library kit (ThermoFisher Scientific, Waltham, USA) was used for the library preparation, and the final library was adjusted to a final concentration of 75 pM using the low TE buffer and loaded onto Ion Chef instrument for emulsion PCR, enrichment, and subsequent placement onto an Ion 540 chip. Next-generation sequencing (NGS) was conducted using the Ion Torrent NGS System using the Ion GenStudio S5 Plus System (Thermo Fisher Scientific, Waltham, USA). Data analysis was performed using Torrent Suite™ software ver.5.18.1. The consensus sequence was analysed using IRMA report ver.1.3.0.2. Annotation was performed using the SnpEff program. The reads were aligned to the *Wuhan Hu-1* strain as a reference genome (NCBI ID: NC_045512.2).

### Phylogenetic and evolutionary analysis of spike protein

The phylogenetic analysis of SARS-CoV-2 was carried out to identify the mutations and relatedness of the JN.1 variant with other SARS-CoV-2 variants using the Nextclade online software v.3.2.0. (https://clades.nextstrain.org/) [10]. The nucleotide sequences coding for spike protein from the datasets available in the Nextclade as reference sequences and reference genomes of SARS-CoV-2 variants from the NCBI database were selected and used to detect the mutations. The Nextclade reference datasets, GenBank accession numbers and the list of mutations are provided in **Supplementary Table 1**. The mutations that were unique to the study sequences are called ‘unique mutations’, whereas those observed in the global JN.1 strains are called ‘universal mutations’. The dynamic mutations characteristically appearing and reappearing in the subsequent variants of SARS-CoV-2 are called ‘dynamic mutations’. Other mutations that were observed with no characteristic pattern were denoted as ‘random mutations’.

### Mutations and superimposition of 3D spike protein structure

Mutations unique to the study sequences were compared with the *Wuhan Hu-1* and other major lineages and sublineages, including XBB and the other reported JN.1 sequences. Mutations were created in the 3D protein structure using PyMol Molecular Graphics System. Using the mutagenesis function and best rotamer based on the frequency of its occurrence and clashes with neighbouring amino acids, the mutation was selected and created. A 3D structure model was created with JN.1 specific mutations using *Wuhan-Hu-1* as the reference (model A), and a model with unique mutations was identified in our study using the reference JN.1 (model B). The above models were superimposed to identify any structural variations due to these mutations. A 3D protein model for the *Wuhan Hu-1* spike protein was built using Swiss-model protein structure homology modelling, upon which mutations were created using the Pymol program. The quality of the developed model was analyzed using SAVES and Procheck online server programs. The root mean square deviation (RMSD) values were used to assess the mutation-induced structural effects on the protein, where a score of 0 indicates identical structures and a high value indicates higher dissimilarity [11, 12].

## RESULTS

### Clinico-demographic characteristics of the JN.1 cohort

The ongoing SARS-CoV-2 genomic surveillance activities at SPHL have identified the dramatic emergence of the JN.1 variant of omicron by replacing the XBB variant between November and December 2023 (**Fig 1A**). The age of the JN.1 positive patients ranged from 1 to 89 years, with the median age during the study period being 51 (**Fig 1B**) and with an equal proportion of male-to-female ratio (**Fig 1C**). More than 50% of the JN.1 patients had diabetes mellitus, followed by 21% of the cases with hypertension, and 10 % of the cases had both diabetes mellitus and hypertension (**Fig 1D**). About 87% of the patients were presenting fever, cold, cough and sore throat (**Fig 1E** & **I**). On analysing the vaccination status of the JN.1 positive patient, 93.5% had received COVID-19 vaccinations, indicating a high proportion of vaccine breakthrough infections (**Fig 1F**). Among the vaccinated, 98% had received two or more doses of COVID-19 vaccines (**Fig 1G**). About 45% of the JN.1 patients were hospitalized, of which 20% had severe illnesses requiring oxygen support and ICU care with ventilator support. However, no deaths were recorded (**Fig 1H**). Among the different symptoms presented by the patients, fever was the most common presentation (69%) and was predominant among the 10-20 years (86%) and 80-90 years (80%) of age. The other common symptoms found were cough (55%) which was predominant in older age groups, rhinorrhea (20%) in younger age groups and sore throat (predominant in 20-40 years (36%) and 60-80 years (32%) age groups. Breathlessness (6%) and myalgia (1.8%) were less prevalent and were mostly seen in older age groups only **(Fig 1I)**.

**FIGURE 1.**
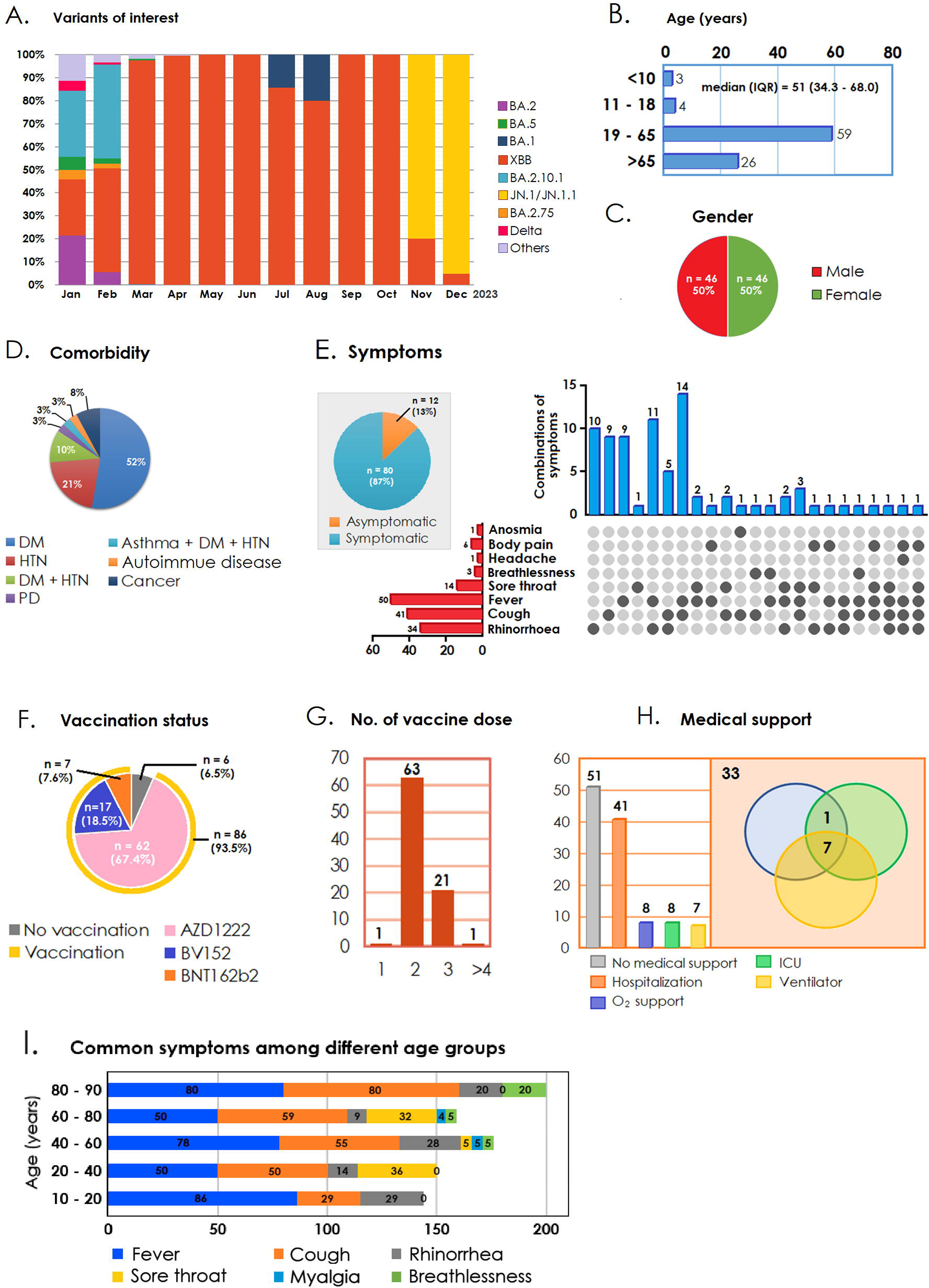
A) Analysis of the proportions of different SARS-CoV-2 variants circulating in Tamil Nadu over time. The x-axis represents the timeline, the y-axis represents the proportion, and different colors represent distinct variants. **B—H)** Clinico-demographic characteristics, where **B)** median age, **C)** gender, **D)** symptoms, **E)** comorbidity, **F)** vaccination status, **G)** number of doses received, **H)** medical support, and **I)** Common symptoms among different age groups. Note: All patients recovered from COVID-19.

### Mutational analysis of spike protein

The S gene sequences of the 66 JN.1 study strains were analysed with different reference genomes and revealed several unique mutations in the different domains of the S protein including the receptor binding domain (RBD), signal peptide (SP) and N-terminal domain (NTD) (**Fig 2** and **Table 1**).

**FIGURE 2.**
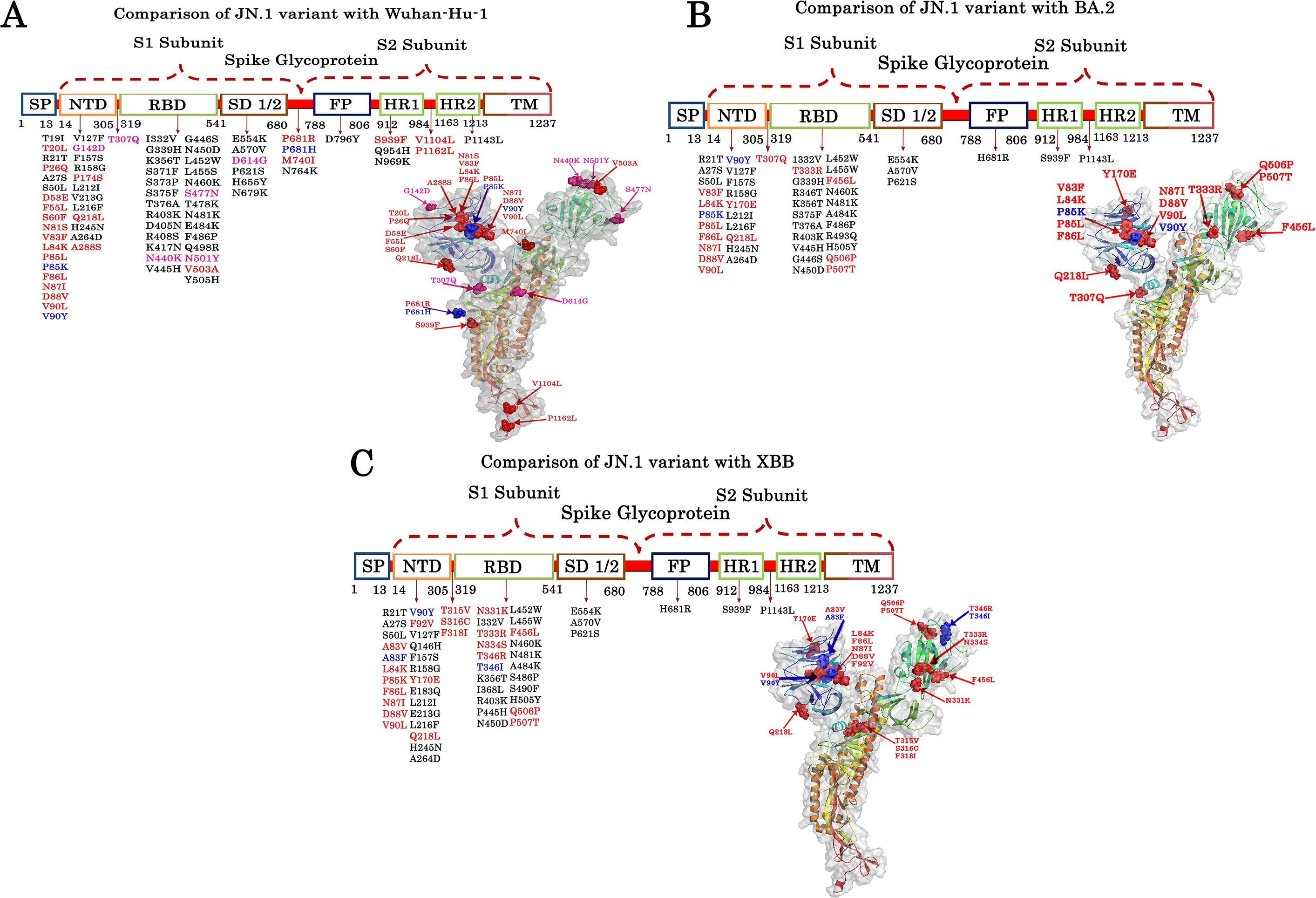
Amino acid changes in the different domains of JN.1 in comparison with *Wuhan-Hu-1*, BA.2, and XBB (Red colour indicates the unique mutations, the blue colour indicates the dynamic mutations, the black colour indicates random mutations and pale red indicates universal mutations). Fusion peptide (FP), Heptad Repeat (HR1/2), N-terminal domain (NTD), Receptor Binding Domain (RBD), Signal peptide (SP), Spike Subdomain (SD) and Transmembrane domain (TM) are the different motifs of the spike glycoprotein.

**Table 1:** Unique and dynamic mutational pattern in spike protein among different lineages.

| S.No | Comparison | Unique Mutations (n) | Unique Mutations | Dynamic Mutations (n) | Dynamic Mutations |
| --- | --- | --- | --- | --- | --- |
| 1 | JN.1 variants with *Wuhan-Hu-1* | 22 | T20L, P26Q, D53E, F55L, S60F, N81S, V83F, L84K, P85L, F86L, N87I, D88V, V90L, P174S, Q218L, A288S, V503A, P681R, M740I, S939F, V1104L, P1162L | 3 | P85K, V90Y, P681R |
| 2 | JN.1 variants with BA.2 | 14 | V83F, L84K, P85L, F86L, N87I, D88V, V90L, Y170E, Q218L, T307Q, T333R, F456L, Q506P, P507T | 2 | P85L, V90Y |
| 3 | JN.1 variants with XBB | 20 | A83V, L84K, P85K, F86L, N87I, D88V, V90L, F92V, Y170E, Q218L, T315V, S316C, F318I, N331K, T333R, N334S, T346R, F456L, Q506P, P507T | 3 | A83F, V90Y, T346I |

### Comparison of spike protein sequences of JN.1 with *Wuhan-Hu-1*

On comparing our JN.1 isolates with the reference sequence of the Wuhan-Hu-1 strain, we have identified 22 unique mutations and 3 dynamic mutations. Of the unique mutations, the majority of them occurred in NTD (n=16), followed by Heptad repeat (HR1/2) (n=3) and spike subdomain (SD) (n=2), whereas RBD showed one mutation (V503A). Among the dynamic mutations, two are seen in NTD (P85K and V90Y) and one in the signal domain (P681H). Also, JN.1 showed six universal mutations (three in NTD, two in RBD-N440K and N501Y and one in SD- D614G). This also encompasses some universal mutations, such as G142D, which have been reported in several countries [13, 14]. Upon analysis of random mutations, we found that JN.1 variants showed 40 mutations across the spike protein when compared to *Wuhan-Hu-1* with a maximum number of mutations in RBD (23) followed by NTD (12) (**Table 2** and **Fig 2A**).

**Table 2:** Unique, dynamic, universal, and random mutations in the spike protein of JN.1 compared to the *Wuhan-Hu-1* sequence.

| UNIQUE MUTATION |  |  |  | DYNAMIC MUTATION |  |  | UNIVERSAL MUTATION |  |  | RANDOM MUTATION |  |  |  |  |
| --- | --- | --- | --- | --- | --- | --- | --- | --- | --- | --- | --- | --- | --- | --- |
| NTD | RBD | SD | HR1/HR2 | NTD | RBD | SD | NTD | RBD | SD | NTD | RBD | SD | FP | HR1/HR2 |
| V83F | V503A | P681R | S939F | P85K | - | P681H | G142D | N440K | D614G | T19I | G339H | N764K | D796Y | Q954H |
| L84K | - | M740I | V1104L | V90Y | - | - | T307Q | S477N | - | R21T | K356T | - | - | N969K |
| P85L | - | - | P1162L | - | - | - | - | N501Y | - | A27S | S371F | - | - | P1143L |
| F86L | - | - | - | - | - | - | - | - | - | S50L | S373P | - | - | - |
| N87I | - | - | - | - | - | - | - | - | - | V127F | S375F | - | - | - |
| D88V | - | - | - | - | - | - | - | - | - | F157S | T376A | - | - | - |
| V90L | - | - | - | - | - | - | - | - | - | R158G | R403K | - | - | - |
| Q218L | - | - | - | - | - | - | - | - | - | L212I | D405N | - | - | - |
| T20L | - | - | - | - | - | - | - | - | - | V213G | R408S | - | - | - |
| P26Q | - | - | - | - | - | - | - | - | - | L216F | K417N | - | - | - |
| D53E | - | - | - | - | - | - | - | - | - | H245N | V445H | - | - | - |
| F55L | - | - | - | - | - | - | - | - | - | A264D | G446S | - | - | - |
| S60F | - | - | - | - | - | - | - | - | - | - | N450D | - | - | - |
| N81S | - | - | - | - | - | - | - | - | - | - | L452W | - | - | - |
| P174S | - | - | - | - | - | - | - | - | - | - | L455S | - | - | - |
| A288S | - | - | - | - | - | - | - | - | - | - | N460K | - | - | - |
| - | - | - | - | - | - | - | - | - | - | - | T478K | - | - | - |
| - | - | - | - | - | - | - | - | - | - | - | N481K | - | - | - |
| - | - | - | - | - | - | - | - | - | - | - | E484K | - | - | - |
| - | - | - | - | - | - | - | - | - | - | - | F486P | - | - | - |
| - | - | - | - | - | - | - | - | - | - | - | Q498R | - | - | - |
| - | - | - | - | - | - | - | - | - | - | - | Y505H | - | - | - |

### Comparison of spike protein sequences of JN.1 with BA.2

Similarly, the results from the comparison of JN.1 with the BA.2 variant show unique mutations, including V83F, L84K, P85L, F86L, N87I, D88V, V90L, Y170E, Q218L, and T307R and dynamic mutations such as P85L, V90Y in the NTD domain and the following unique mutations T333R, F456L, Q506P, and P507T found within the RBD domain. There was no occurrence of universal mutations within any of the domains. This shows the amino acid change compared to BA.2 were identified individually (**Table 3** and **Fig 2B**). We also found a total of 34 random mutations when comparing JN.1 and BA.2 with 10 mutations in NTD and 18 in RBD. The fusion peptide (FP) domain showed a mutation – H681R.

**Table 3:** Unique, dynamic, universal, and random mutations in the spike protein of JN.1 compared to the BA.2 sequence.

| UNIQUE MUTATION |  |  |  | DYNAMIC MUTATION |  |  | UNIVERSAL MUTATION |  |  | RANDOM MUTATION |  |  |  |  |
| --- | --- | --- | --- | --- | --- | --- | --- | --- | --- | --- | --- | --- | --- | --- |
| NTD | RBD | SD | HR1/HR2 | NTD | RBD | SD | NTD | RBD | SD | NTD | RBD | SD | FP | HR1/HR2 |
| V83F | T33R | - | - | P85K | - | - | - | - | - | R21T | I332V | E554K | H681R | S939F |
| L84K | F456L | - | - | V90Y | - | - | - | - | - | A27S | G339H | A570V | - | P1143L |
| P85L | Q506P | - | - | - | - | - | - | - | - | S50L | R346T | P621S | - |  |
| F86L | P507T | - | - | - | - | - | - | - | - | V127F | K356T | - | - | - |
| N87I | - | - | - | - | - | - | - | - | - | F157S | S375F | - | - | - |
| D88V | - | - | - | - | - | - | - | - | - | R158G | T376A | - | - | - |
| V90L | - | - | - | - | - | - | - | - | - | L212I | R403K | - | - | - |
| Q218L | - | - | - | - | - | - | - | - | - | L216F | V445H | - | - | - |
| Y170E | - | - | - | - | - | - | - | - | - | H245N | G446S | - | - | - |
| T307Q | - | - | - | - | - | - | - | - | - | A264D | N450D | - | - | - |
| - | - | - | - | - | - | - | - | - | - | - | L452W | - | - | - |
| - | - | - | - | - | - | - | - | - | - | - | L455W | - | - | - |
| - | - | - | - | - | - | - | - | - | - | - | N460K | - | - | - |
| - | - | - | - | - | - | - | - | - | - | - | N481K | - | - | - |
| - | - | - | - | - | - | - | - | - | - | - | A484K | - | - | - |
| - | - | - | - | - | - | - | - | - | - | - | F486P | - | - | - |
| - | - | - | - | - | - | - | - | - | - | - | R493Q | - | - | - |
| - | - | - | - | - | - | - | - | - | - | - | H505Y | - | - | - |

### Comparison of spike protein sequences of JN.1 with XBB

Likewise, the comparison of JN.1 with the XBB variant showed no traces of universal mutations and was found to have unique, dynamic, and random mutations. The unique mutations in the NTD domain include A83V, L84K, P85K, F86L, N87I, D88V, V90L, F92V, Y170E, and Q218L followed had two dynamic mutations (A83F and, V90Y). In the RBD domain, unique mutations, including N331K, T333R, N334S, T346R, F456L, Q507P, and P507T and dynamic mutation-T346I were identified. The other unique mutations, T315V, S316C and F318I, were spotted in the intermediary region between NTD and RBD domains (**Table 4** and Figure 2C). Our study shows 33 random mutations (13 in NTD, 14 in RBD, three in SD1/2, two in HR1-2 and one in the FP).

**Table 4:** Unique, dynamic, universal, and random mutations in the spike protein of JN.1 compared to the XBB sequence.

| UNIQUE MUTATION |  |  |  | DYNAMIC MUTATION |  |  | UNIVERSAL MUTATION |  |  | RANDOM MUTATION |  |  |  |  |
| --- | --- | --- | --- | --- | --- | --- | --- | --- | --- | --- | --- | --- | --- | --- |
| NTD | RBD | SD | HR1/HR2 | NTD | RBD | SD | NTD | RBD | SD | NTD | RBD | SD | FP | HR1/HR2 |
| A83V | N331K | - | - | A83F | T346I | - | - | - | - | R21T | I332V | E554K | H681R | S939F |
| L84K | T333R | - | - | V90Y | - | - | - | - | - | A27S | K356T | A570V | - | P1143L |
| P85K | N334S | - | - | - | - | - | - | - | - | S50L | I368L | P621S | - | - |
| F86L | T346R | - | - | - | - | - | - | - | - | V127F | R403K | - | - | - |
| N87I | F456L | - | - | - | - | - | - | - | - | Q146H | P445H | - | - | - |
| D88V | Q506P | - | - | - | - | - | - | - | - | F157S | N450D | - | - | - |
| V90L | P507T | - | - | - | - | - | - | - | - | R158G | L452W | - | - | - |
| F92V | - | - | - | - | - | - | - | - | - | E183Q | L455W | - | - | - |
| Y170E | - | - | - | - | - | - | - | - | - | L212I | N460K | - | - | - |
| Q218L | - | - | - | - | - | - | - | - | - | E213G | N481K | - | - | - |
| T315V | - | - | - | - | - | - | - | - | - | L216F | A484K | - | - | - |
| S316C | - | - | - | - | - | - | - | - | - | H245N | S486P | - | - | - |
| F318I | - | - | - | - | - | - | - | - | - | A264D | S490F | - | - | - |
| - | - | - | - | - | - | - | - | - | - | - | H505Y | - | - | - |

### Evolutionary mutation patterns in spike protein sequences

Furthermore, the evolutionary relatedness of JN.1 is presented in a phylogenetic tree that depicts the hierarchy of the different lineages of the selected strain and the position of the JN.1 lineage in the tree. The JN.1 lineage showed an extended branch length in both rooted and unrooted trees (**Fig 3**).

**FIGURE 3.**
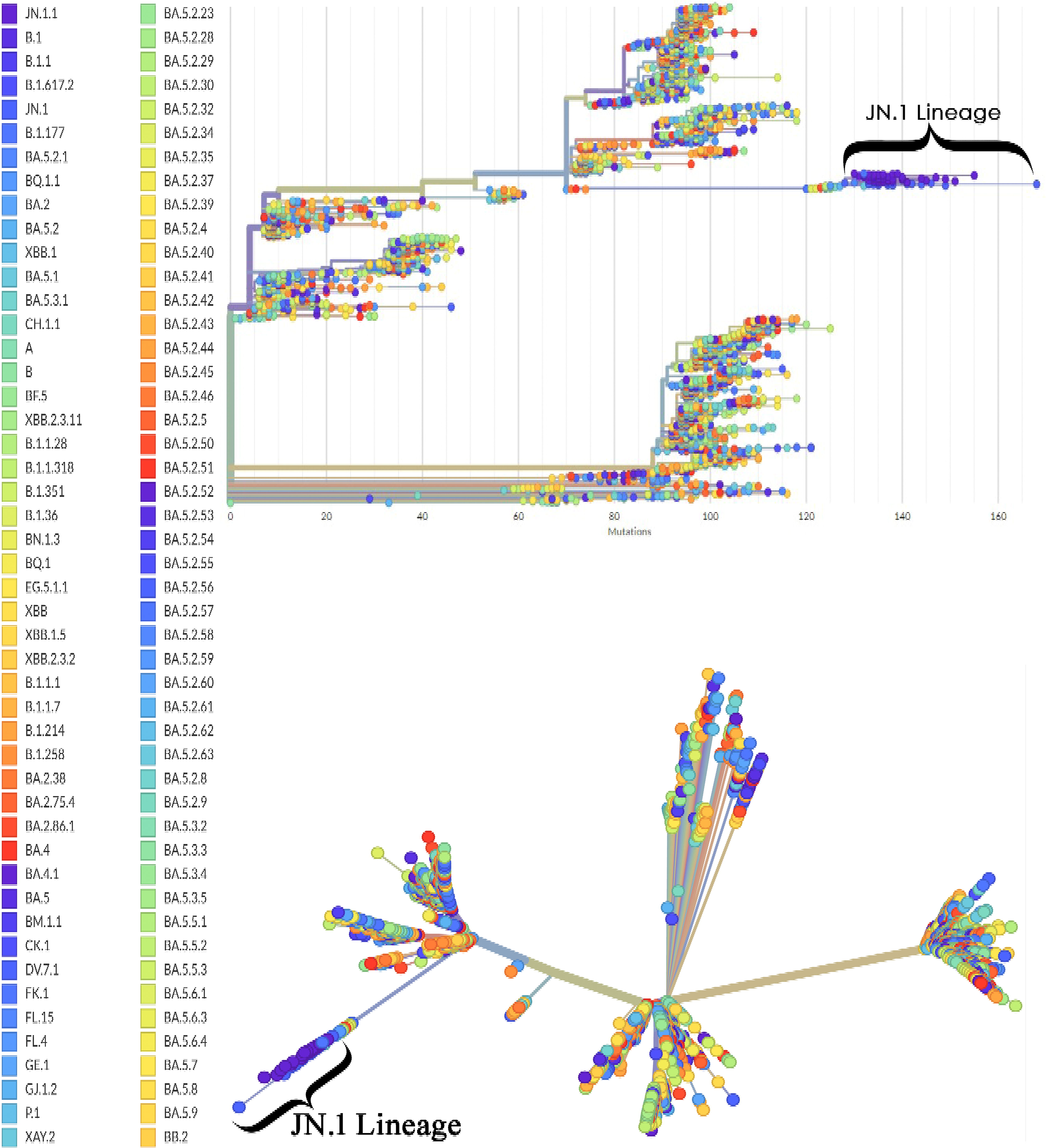
The phylogenetic analysis of the JN.1 variant using a rooted tree and an unrooted tree illustrates the evolutionary relatedness among the sequences of JN.1 (n=66) compared with the reference genome of SARS-CoV-2 (*Wuhan-Hu-1*).

### Mutational and conformational changes in spike protein

When our study sequences were compared with the reference genomes of different variants, 19 mutations were observed in our strains and previously reported JN.1 variants (but not seen in other lineages). This indicated that the mutations were unique to the JN.1 variant. The list of mutations and the specific motifs is given in **Table 5**, and the frequency of these mutations is given in **Supplementary Table 1**. The other 24 mutations that were observed in a few lineages/sub-lineages but not found in our 66 sequences were identified and listed in **Table 6**.

**Table 5:** List of mutations observed in our strains and previously reported JN.1 variants.

| S. no | Mutations | Motif | Subunit |
| --- | --- | --- | --- |
| 1 | R21T | NTD | S1 |
| 2 | S50L | NTD |  |
| 3 | V127F | NTD |  |
| 4 | F157S | NTD |  |
| 5 | R158G | NTD |  |
| 6 | L216F | NTD |  |
| 7 | H245N | NTD |  |
| 8 | A270D | NTD |  |
| 9 | I332V | RBD |  |
| 10 | K356T | RBD |  |
| 11 | R403K | RBD |  |
| 12 | N450D | RBD |  |
| 13 | L455S | RBD |  |
| 14 | N481K | RBD |  |
| 15 | E554K | SD1/2 |  |
| 16 | A570V | SD1/2 |  |
| 17 | P621S | SD1/2 |  |
| 18 | S939F | HR1 | S2 |
| 19 | P1143L | HR2 |  |

**Table 6:** List of mutations observed in a few lineages/sub-lineages but not in our study.

| **S. No** | **Mutations** | **Motif** | **Variants bearing the mutation** |
| --- | --- | --- | --- |
| 1. | L24S | NTD | BA.2; BA.2.75; BA.2.10; BA.4; BA.5 |
| 2. | A67V | NTD | BA.1 |
| 3. | H146Q | NTD | XBB.1 |
| 4. | K147E | NTD | BA.2.75 |
| 5. | W152R | NTD | BA.2.75 |
| 6. | F157L | NTD | BA.2.75 |
| 7. | Q183E | NTD | XBB1 |
| 8. | I210V | NTD | BA.2.75 |
| 9. | L212V | NTD | BA.1 |
| 10. | R214K | NTD | BA.1 |
| 11. | G252V | NTD | BA.1 |
| 12. | G257S | NTD | BA.2.75 |
| 13. | L368I | RBD | XBB1 |
| 14. | Y449S | RBD | BA.1 |
| 15. | F490L | RBD | BA.1 |
| 16. | Q493R | RBD | BA.2;BA.1.1.529;BA2.10 |
| 17. | G496N | RBD | BA.1 |
| 18. | T500A | RBD | BA.1 |
| 19. | G502S | RBD | XBB1 |
| 20. | N658S | SD1/2 | BA.4 |
| 21. | Q755R | SD1/2-FP | BA.2.75 |
| 22. | D950N | HR1 | BA.1.617.2 |
| 23. | L981F | HR1 | BA.1 |
| 24. | S982A | HR1 | BA.1.1 |
| 25. | V1228L | TM | BA.1 |

Intrigued by these mutations, we next elucidated the resulting protein structural changes with the reference protein sequences. The 19 JN.1-specific mutations were created in the model and superimposed with the reference protein. The superimposed structure showed an RMSD value of 0.071Å (**Fig 4A**). The 17 study-specific mutations were created and superimposed with the same model, which showed an RMSD value of 0.081Å (**Fig 4B**).

**FIGURE 4.**
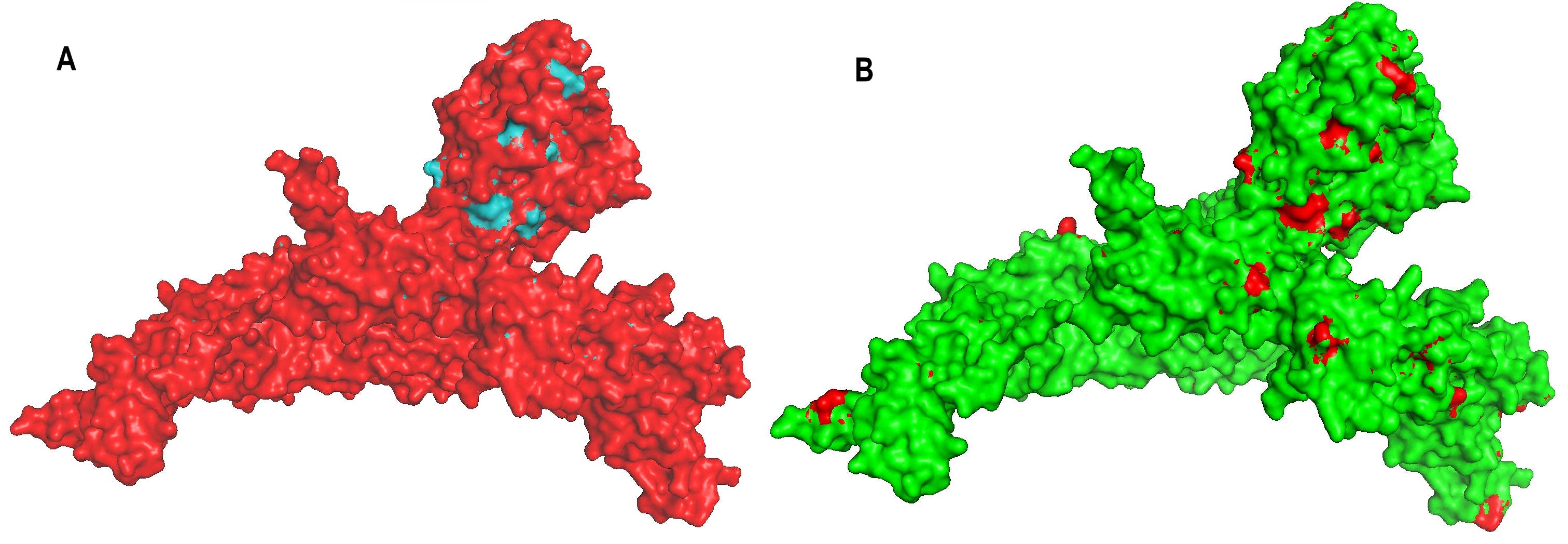
Superimposition of protein models. **A:** Superimposed protein model of *Wuhan Hu-1* reference model (red) with JN.1 specific mutations (blue). **B:** Superimposed protein model of JN.1 variant (green) with the study-specific unique mutation model identified in our study patients (red).

## DISCUSSION

Here, the study describes the detection of SARS-CoV-2 Omicron subvariant JN.1 during November and December-2023, as a part of the state public health genomic surveillance activity which has been happening since September 2021. In our recent study, we showed the mutational patterns of Omicron variants and the emergence of the XBB as a dominant variant in January 2023 replacing BA.2 which continued till October 2023 [15, 16]. In September 2023, JN.1 was first identified in the United States [17] followingly many countries like Canada, France, Singapore and the United Kingdom reported the emergence of JN.1 [18–21]. In India, the JN.1 variant was first identified on 6th October 2023. In Tamil Nadu, the first case of JN.1 was reported on 27 December 2023, subsequently since then, JN.1 replaced the XBB variant and was the most predominant SARS-CoV-2 variant in the state of Tamil Nadu, India.

Along with the L455S, a hallmark FLip mutation of JN.1 in the spike protein, the present study identified mutations in different domains of the protein, particularly in the NTD, RBD, SD1/2 and HR1/2 domains. Furthermore, the study identified several mutations when compared to Wuhan-Hu-1, BA.2 and XBB. This indicates the adaptive evolutionary trends of the virus possibly due to reduced neutralizing antibody responses [22, 23].

Since the emergence of JN.1 in August 2023 as a descendant of the BA.2.86 lineage, it drew much attention due to mutation-induced interference in viral binding to angiotensin-binding enzyme (ACE2) receptors [24, 25]. A significant rise in numbers of JN.1 variant, post vaccinations indicates breakthrough infections with enhanced capacity of immune evasion [26]. The predecessor XBB variant had 82% of immune evasiveness among the vaccinated individuals [15], whereas the current JN.1 variant by acquiring several mutations led to structural changes in the spike protein, conferring 95% of immune evasiveness [27]. This warranted a detailed investigation on other additional mutations that JN.1 harboured subsequently from its predecessors [28]. The present study analyzed the mutational dynamics of the virus it inherited across the lineages and sublineages. Though the study does not represent the true prevalence of the JN.1 in India, the widespread distribution of this variant in the community warrants nationwide genomic surveillance of SARS-CoV-2 under the INSACOG network.

Although viruses are constantly known to evolve due to genetic diversity and evolutionary selection pressure, the clinical and immunological outcomes are always unpredictable. It is therefore important to carry out continuous surveillance on SARS-CoV-2 variants and their mutational analysis [29]. The mutational pattern leads to virus evolution resulting in the emergence of variants with enhanced transmissibility, severity and immune evasion [30]. The disappearance and re-appearance of certain mutations (dynamic mutations), in the course of virus evolution with any possible clinical relevance is generally overlooked in evolutionary studies. The present study takes cognizance of such dynamic patterns during virus evolution. The analysis revealed dynamic mutations such as P85L to P85K, V90L to V90Y in the NTD domain and P681R to P681H in the intermediate region between SD1/2 and FP. These dynamic mutations were identified by comparing JN.1 with the *Wuhan-Hu-1*, BA.2 and XBB. The mutational spectrum in the NTD domain of the spike protein has been an important mechanism in driving antigenic variation and host adaptation [31]. Studies indicate that potent neutralizing antibodies to NTD interact with residues F140S, G142D, Y145D, K150E, W152R and R158S [32]. The study also showed that antibodies against both RBD and NTD could efficiently neutralize SARS-CoV-2, limiting the emergence of neutralization-escape mutants.

Our study indicated universal mutation-G142D and common mutations-Q146H and R158G identified in JN.1 variants in line with other studies [32, 33]. These findings further augment the emergence of vaccine-escape mutants. Similarly, by comparing the JN.1 with XBB, we show two dynamic mutations - A83V to A83F, V90L to V90Y in the NTD domain and T346R to T346I in the RBD domain. This indicates that these mutations would possibly contribute to the ongoing evolution of viral lineages, and their overall fitness and adaptability [34] Our study identified a dynamic mutation P85K that has gradually evolved from the *Wuhan-Hu-1* and BA.2 strain and reshaped into a unique mutation in comparison with XBB [15]. This indicates the genetic drift within the SARS-CoV-2 infection particularly in the JN.1 variant. These results were further supported by the phylogenetic tree where the JN.1 lineage showed an increased branch length demonstrating both the evolutionary divergence and its relationships with the other lineages of SARS-CoV-2 [35].

The impact of unique mutations identified in the spike protein of the study strains on its structure was analyzed by superimposing the mutant model with the wild-type model. The structural alignment between the two models was analyzed with the equivalent backbone atoms. Every single mutation causes concomitant changes in protein folding, conformation, physiochemical properties and *in situ* function [36]. The conformational changes upon superimposition are analyzed with RMSD as a measurement. The predicted structural modifications on ab initio protein models are a known limitation on the robustness yet considered to be the best approach for comprehensive analysis of variants [7]. Protein function predictions are now studied increasingly through computational methods as the curated protein structures, along with experimentally determined host-pathogen protein interactions [8]. In the present study, as anticipated, the superimposed structure showed only a slight deviation (<0.1Å) compared to the reference strains. Nevertheless, these mutations may be the prelude for further mutations in the course of the evolution of the new sub-lineage. Therefore, monitoring the periodical changes in the virus dynamics is important [37]. Through the combination of high-throughput sequencing technologies with phylogenetic analysis, it is possible to assess parallel patterns of evolution driving significant phenotypic shifts. These methods offer a framework to measure and predict future evolutionary occurrences [38].

The present study identified 19 unique mutations among the study sequences. The majority of the mutations occurred in the NTD and RBD domains followed by the HR1/2 region. The mutations and the following structural changes in the NTD have been implicated in reduced epitope recognition ensuing immune viral escape. The mutations of the RBD domain have been implicated significantly with infectability, transmissibility, and antibody resistance. Spike proteins are known to accumulate multiple mutations upon evolution, subsequently ranking up the virion spike density and infectivity. The present study identified novel mutations of the JN.1 variant, possibly contributing to high transmissibility and immune resistance. In addition, the structural conformation analysis by superimposing them with reference protein structure indicated structural divergence. The considerable deviations measured in conventional RMSD could have been studied further to elucidate its functional variability. Our study also identified 24 mutations that were present in previous lineages but lost in the JN.1 variant, indicating the dynamicity of such mutations. These dynamic mutations might play a substantial role in the viral evolution and pathogenicity mechanism and are to be studied.

## CONCLUSIONS

Recognizing the significance of mutations is crucial for modifying public health strategies and therapeutic interventions. This study aimed to characterize the clinical profiling and genetic modifications of the JN.1 variant to the previous variants of SARS-CoV-2, including the wild type *Wuhan-Hu-1*, BA.2, and XBB. The study showed that the prevalent mutations in JN.1 may be responsible for their immune evasive properties, and acknowledged that the genetic landscape of SARS-CoV-2 is inconstant and this capability to identify and respond to alterations in the genetic composition of the virus is vital for effective pandemic governance.

## Author contributions

Sivaprakasam T. Selvavinayagam, Masilamani Senthilkumar, Yean K. Yong, Suvaiyarasan Suvaithenamudhan, Amudhan Murugesan, Marie Larsson, Pachamuthu Balakrishnan, Siddappa N. Byrareddy, Vijayakumar Velu, Esaki M. Shankar, and Sivadoss Raju designed the study and were responsible for conceptualization and data curation. Sivaprakasam T. Selvavinayagam, Narayanaiah Cheedarla, Kannan Hemashree, Sathish Sankar, Yean K. Yong, Hong Y. Tan, Ying Zhang, Parthiban Arthydevi, Ramendra P. Pandey, Manivannan Rajeshkumar, Anandhazhvar Kumaresan, Natarajan Gopalan, Meganathan Kannan, Marie Larsson, Saravanan Shanmugam, Vijayakumar Velu, Pachamuthu Balakrishnan, Esaki M.Shankar, and Sivadoss Raju conducted the analysis, and were responsible for methodology, formal analysis, validation, and visualization. Sivaprakasam T. Selvavinayagam, Yean K. Yong, Marie Larsson, Vijayakumar Velu, Esaki M. Shankar, and Sivadoss Raju wrote the first draft of the manuscript. All authors provided critical inputs and approved the final version of the manuscript for publication.

## Ethical approval statement

The study was approved by the Institutional Ethical Committee of Madras Medical College and Hospital, Chennai (EC No. 03092021).

## Patient consent statement

Patient written informed consent was obtained.

## Acknowledgements

S.T.S. and S.R. are funded by the National Health Mission, Tamil Nadu (680/NGS/NHMTNMSC/ENGG/2021) for the Directorate of Public Health and Preventive Medicine, WGS facility. M.L. is supported by grants through AI52731, the Swedish Research, the Swedish Physicians against AIDS Research Foundation, the Swedish International Development Cooperation Agency, SIDASARC, VINNMER for Vinnova, Linköping University Research Fund, CALF, and the Swedish Society of Medicine. V.V. is supported by the Office of Research Infrastructure Programs(ORIP/NIH) base grant P51 OD011132 to ENPRC. A.M. is supported by Grant No. 12020/04/2018LJHR, Department of Health Research, Government of India. The funders of this study had no role in the study design, data collection, data analysis, data interpretation, or writing of the report. The authors thank the Indian SARS CoVLJ2 Genomic Consortium (INSACOG), Department of Biotechnology, Ministry of Science and Technology, Government of India for their approval and inclusion of the State Public Health Laboratory (SPHL) as INSAGOG Genomic Sequencing Laboratory (IGSL) vide File No: RADLJ22017/28/2020LJKGDDBTLJPart (6) Dated 29th December 2021.

## Data Availability Statement

The data that support the findings of this study are available from the corresponding author upon reasonable request. The identified clinical data of patients in the study and the ‘in-house’ bioinformatics pipeline in Python are available on request to the corresponding authors.

## Abbreviations

ACE2, angiotensinLJconverting enzyme-2; CDC, Centers for Disease Control and Prevention; cDNA, complementary DNA; COVIDLJ19, coronavirus disease 2019; Ct, cycle threshold value; FP, fusion peptide; HDU, high dependency unit; HR, heptad repeat; ICU, intensive care unit; Polymerase Chain Reaction; NGS, nextLJgeneration sequencing; NTD, NLJterminal domain; QC, quality control; RBD, receptor binding domain; RMSD, root mean square deviation; RT-PCR, Reverse transcription PCR; SARS-CoV-2, Severe Acute Respiratory Syndrome Coronavirus 2; SD, spike subdomain; SP: signal peptide; SPHL, State Public Health Laboratory; TE, Tris-EDTA; TM: Transmembrane domain; VOC, variants of concern; WGS, whole genome sequencing.

## References

1. Zaidi AK, Singh RB. SARS-CoV-2 variant biology and immune evasion. Prog Mol Biol Transl Sci 2024;202:45–66.

2. Fan Y, Li X, Zhang L, Wan S, Zhang L, et al. SARS-CoV-2 Omicron variant: recent progress and future perspectives. Signal Transduction and Targeted Therapy 2022 7:1 2022;7:1–11.

3. Chakraborty C, Bhattacharya M, Chopra H, Islam MA, Saikumar G, et al. The SARS-CoV-2 Omicron recombinant subvariants XBB, XBB.1, and XBB.1.5 are expanding rapidly with unique mutations, antibody evasion, and immune escape properties – an alarming global threat of a surge in COVID-19 cases again? Int J Surg 2023;109:1041.

4. Selvavinayagam ST, Yong YK, Joseph N, Hemashree K, Tan HY, et al. Low SARS-CoV-2 viral load among vaccinated individuals infected with Delta B.1.617.2 and Omicron BA.1.1.529 but not with Omicron BA.1.1 and BA.2 variants. Front Public Health 2022;10:1018399.

5. Mahilkar S, Agrawal S, Chaudhary S, Parikh S, Sonkar SC, et al. SARS-CoV-2 variants: Impact on biological and clinical outcome. Front Med (Lausanne);9. Epub ahead of print 10 November 2022. DOI: 10.3389/FMED.2022.995960.

6. Kumari M, Lu RM, Li MC, Huang JL, Hsu FF, et al. A critical overview of current progress for COVID-19: development of vaccines, antiviral drugs, and therapeutic antibodies. J Biomed Sci;29. Epub ahead of print 1 December 2022. DOI: 10.1186/S12929-022-00852-9.

7. Lee D, Redfern O, Orengo C. Predicting protein function from sequence and structure. Nat Rev Mol Cell Biol 2007;8:995–1005.

8. Pál C, Papp B, Lercher MJ. An integrated view of protein evolution. Nat Rev Genet 2006;7:337– 348.

9. Lomoio U, Puccio B, Tradigo G, Guzzi PH, Veltri P. SARS-CoV-2 protein structure and sequence mutations: Evolutionary analysis and effects on virus variants. PLoS One 2023;18:e0283400.

10. Aksamentov I, Roemer C, Hodcroft E, Neher R. Nextclade: clade assignment, mutation calling and quality control for viral genomes. J Open Source Softw 2021;6:3773.

11. Ravantti JJ, Martinez-Castillo A, Abrescia NGA. Superimposition of Viral Protein Structures: A Means to Decipher the Phylogenies of Viruses. Viruses;12. Epub ahead of print 1 October 2020. DOI: 10.3390/V12101146.

12. Carugo O, Pongor S. A normalized root-mean-spuare distance for comparing protein three- dimensional structures. Protein Sci 2001;10:1470.

13. Davis JJ, Long SW, Christensen PA, Olsen RJ, Olson R, et al. Analysis of the ARTIC Version 3 and Version 4 SARS-CoV-2 Primers and Their Impact on the Detection of the G142D Amino Acid Substitution in the Spike Protein. Microbiol Spectr;9. Epub ahead of print 22 December 2021. DOI: 10.1128/SPECTRUM.01803-21.

14. Singh P, Sharma K, Singh P, Bhargava A, Negi SS, et al. Genomic characterization unravelling the causative role of SARS-CoV-2 Delta variant of lineage B.1.617.2 in 2nd wave of COVID-19 pandemic in Chhattisgarh, India. Microb Pathog;164. Epub ahead of print 1 March 2022. DOI: 10.1016/J.MICPATH.2022.105404.

15. Selvavinayagam ST, Karishma SJ, Hemashree K, Yong YK, Suvaithenamudhan S, et al. Clinical characteristics and novel mutations of omicron subvariant XBB in Tamil Nadu, India - a cohort study. The Lancet regional health Southeast Asia;19. Epub ahead of print 1 December 2023. DOI: 10.1016/J.LANSEA.2023.100272.

16. Selvavinayagam ST, Suvaithenamudhan S, Yong YK, Hemashree K, Rajeshkumar M, et al. Genomic surveillance of omicron B.1.1.529 SARS-CoV-2 and its variants between December 2021 and March 2023 in Tamil Nadu, India—A state-wide prospective longitudinal study. J Med Virol 2024;96:e29456.

17. Looi MK. Covid-19: WHO adds JN.1 as new variant of interest. BMJ 2023;383:p2975.

18. Planas D, Staropoli I, Michel V, Lemoine F, Donati F, et al. Distinct evolution of SARS-CoV-2 Omicron XBB and BA.2.86/JN.1 lineages combining increased fitness and antibody evasion. Nat Commun;15. Epub ahead of print 1 December 2024. DOI: 10.1038/S41467-024-46490-7.

19. Wannigama DL, Amarasiri M, Phattharapornjaroen P, Hurst C, Modchang C, et al. Wastewater-based epidemiological surveillance of SARS-CoV-2 new variants BA.2.86 and offspring JN.1 in south and Southeast Asia. J Travel Med. Epub ahead of print 4 March 2024. DOI: 10.1093/JTM/TAAE040.

20. Ou G, Yang Y, Zhang S, Niu S, Cai Q, et al. Evolving immune evasion and transmissibility of SARS-CoV-2: The emergence of JN.1 variant and its global impact. Drug Discov Ther;18. Epub ahead of print 29 February 2024. DOI: 10.5582/DDT.2024.01008.

21. Kaku Y, Okumura K, Padilla-Blanco M, Kosugi Y, Uriu K, et al. Virological characteristics of the SARS-CoV-2 JN.1 variant. Lancet Infect Dis 2024;24:e82.

22. Wang Q, Iketani S, Li Z, Liu L, Guo Y, et al. Alarming antibody evasion properties of rising SARS-CoV-2 BQ and XBB subvariants. Cell 2023;186:279–286.e278.

23. Patel N, Trost JF, Guebre-Xabier M, Zhou H, Norton J, et al. XBB.1.5 spike protein COVID-19 vaccine induces broadly neutralizing and cellular immune responses against EG.5.1 and emerging XBB variants. Scientific Reports 2023 13:1 2023;13:1–11.

24. Kaku Y, Okumura K, Padilla-Blanco M, Kosugi Y, Uriu K, et al. Virological characteristics of the SARS-CoV-2 JN.1 variant. Lancet Infect Dis 2024;24:e82.

25. Uriu K, Ito J, Kosugi Y, Tanaka YL, Mugita Y, et al. Transmissibility, infectivity, and immune evasion of the SARS-CoV-2 BA.2.86 variant. Lancet Infect Dis 2023;23:e460–e461.

26. Yang S, Yu Y, Xu Y, Jian F, Song W, et al. Fast evolution of SARS-CoV-2 BA.2.86 to JN.1 under heavy immune pressure. Lancet Infect Dis 2024;24:e70–e72.

27. Karyakarte RP, Das R, Rajmane M V, Dudhate S, Agarasen J, et al. Appearance and Prevalence of JN.1 SARS-CoV-2 Variant in India and Its Clinical Profile in the State of Maharashtra. Cureus. Epub ahead of print 22 March 2024. DOI: 10.7759/cureus.56718.

28. Kosugi Y, Plianchaisuk A, Putri O, Uriu K, Kaku Y, et al. Characteristics of the SARS-CoV-2 omicron HK.3 variant harbouring the FLip substitution. Lancet Microbe;0. Epub ahead of print January 2024. DOI: 10.1016/s2666-5247(23)00373-7.

29. Singh J, Pandit P, McArthur AG, Banerjee A, Mossman K. Evolutionary trajectory of SARS-CoV- 2 and emerging variants. Virology Journal 2021 18:1 2021;18:1–21.

30. Goga A, Bekker LG, Garrett N, Reddy T, Yende-Zuma N, et al. Breakthrough SARS-CoV-2 infections during periods of delta and omicron predominance, South Africa. The Lancet 2022;400:269–271.

31. Cantoni D, Murray MJ, Kalemera MD, Dicken SJ, Stejskal L, et al. Evolutionary remodelling of N-terminal domain loops fine-tunes SARS-CoV -2 spike . EMBO Rep;23. Epub ahead of print 6 October 2022. DOI: 10.15252/EMBR.202154322/SUPPL_FILE/EMBR202154322-SUP-0002-DATASETEV1.XLSX.

32. Haslwanter D, Dieterle ME, Wec AZ, O’brien CM, Sakharkar M, et al. A Combination of Receptor-Binding Domain and N-Terminal Domain Neutralizing Antibodies Limits the Generation of SARS-CoV-2 Spike Neutralization-Escape Mutants. mBio;12. Epub ahead of print 1 October 2021. DOI: 10.1128/MBIO.02473-21.

33. Suryadevara N, Shrihari S, Gilchuk P, VanBlargan LA, Binshtein E, et al. Neutralizing and protective human monoclonal antibodies recognizing the N-terminal domain of the SARS-CoV- 2 spike protein. Cell 2021;184:2316–2331.e15.

34. Harvey WT, Carabelli AM, Jackson B, Gupta RK, Thomson EC, et al. SARS-CoV-2 variants, spike mutations and immune escape. Nat Rev Microbiol 2021;19:409–424.

35. Negi SS, Schein CH, Braun W. Regional and temporal coordinated mutation patterns in SARS- CoV-2 spike protein revealed by a clustering and network analysis. Sci Rep;12. Epub ahead of print 1 December 2022. DOI: 10.1038/S41598-022-04950-4.

36. Ravantti JJ, Martinez-Castillo A, Abrescia NGA. Superimposition of Viral Protein Structures: A Means to Decipher the Phylogenies of Viruses. Viruses;12. Epub ahead of print 1 October 2020. DOI: 10.3390/V12101146.

37. Bloom JD, Beichman AC, Neher RA, Harris K. Evolution of the SARS-CoV-2 Mutational Spectrum. Mol Biol Evol;40. Epub ahead of print 4 April 2023. DOI: 10.1093/MOLBEV/MSAD085.

38. Dolan PT, Whitfield ZJ, Andino R. Mapping the Evolutionary Potential of RNA Viruses. Cell Host Microbe 2018;23:435–446.

